# Estimating Causal Treatment Effects of Femoral and Tibial Derotational Osteotomies on Foot Progression in Children with Cerebral Palsy

**DOI:** 10.1101/2021.03.04.21252476

**Authors:** Michael H. Schwartz, Hans Kainz, Andrew G. Georgiadis

## Abstract

**Background:** Foot progression deviations are a common and important problem for children with CP. Tibial and femoral derotational osteotomies (TDO, FDO) are used to treat foot progression deviations, but outcomes are unpredictable. The available evidence for the causal effects of TDO and FDO is limited and weak, and thus modeling approaches are needed.

**Methods:** We queried our clinical database for individuals with a diagnosis of cerebral palsy (CP) who were less than 18 years old and had baseline and follow up gait data collected within a 3-year time span. We then used the Bayesian Causal Forest (BCF) algorithm to estimate the causal treatment effects of TDO and FDO on foot progression deviations (separate models). We examined average treatment effects and heterogeneous treatment effects (HTEs) with respect to clinically relevant covariates.

**Results:** The TDO and FDO models were able to accurately predict follow-up foot progression (r^2^ ∼0.7, RMSE ∼8°). The estimated causal effect of TDO was bimodal and exhibited significant heterogeneity with respect to baseline levels of foot progression and tibial torsion as well as changes in tibial torsion at follow-up. The estimated causal effect of FDO was unimodal and largely homogeneous with respect to baseline or change characteristics.

**Conclusions:** This study demonstrated the potential for causal machine-learning algorithms to impact treatment in children with CP. The causal model is accurate and appears sensible – though no gold-standard exists for validating the causal estimates. The model results can provide guidance for planning surgical corrections, and partly explain unsatisfactory outcomes observed in prior observational clinical studies.

## 1 Background

The primary goal of this study is to develop methods that will improve short-term outcomes for derotational osteotomies intended to correct foot progression deviations in children with cerebral palsy (CP). To do this, we first estimate the causal treatment effects of tibial and femoral derotational osteotomies (TDO and FDO, respectively) on foot progression. We then investigate heterogeneous treatment effects (HTE) to identify preoperative patient characteristics that influence outcomes. The secondary goal of the study is to demonstrate the utility of the Bayesian Causal Forest (BCF) algorithm for analyzing treatments in this complex patient population where strong evidence, such as results of randomized controlled trials (RCT), is rare or non-existent.

### 1.1 Importance of Foot Progression

Foot progression deviations are a common and important problem in CP. Internal foot progression deviations are often felt to be cosmetically undesirable. External foot progression associated with external tibial torsion has been shown to produce biomechanical deficits that could lead to crouch gait (1). The mechanical loading of an externally or internally deviated foot is atypical, and thus believed to be a contributing factor to progressive foot deformity (2). Finally, foot progression deviations can cause atypical loading of the knee joint in the coronal and transverse planes, which could, over time, hasten or worsen degenerative knee conditions such as osteoarthritis (3,4).

### 1.2 Tibial and Femoral Derotational Osteotomies

Tibial torsion and femoral anteversion deformities are widely believed to contribute to foot progression deviations. However, careful radiographic analysis by Lee et al., showed that tibial torsion and femoral anteversion only explained 18% of the variance in foot progression deviation (5). Westberry et al., showed poor correlations between radiographic measures of femoral anteversion, physical exam based measures of anteversion, and dynamic presentation of internal rotation gait (6).

Tibial and femoral derotational osteotomies are common orthopedic procedures used to correct foot progression deviations. Both procedures have been studied extensively, but generally using observational and non-causal designs. Kay et al. showed that FDOs produced dynamic corrections that were 40% smaller than the operative derotation (7). Braatz et al. later showed no correlation between femoral anteversion and mean hip rotation during the stance phase of gait, and concluded that outcomes of FDOs will be unsatisfactory if the magnitude of operative derotation is based on anteversion angle (8). In contrast, the response to TDOs appears more predictable. Ryan et al. showed that the amount of change in foot progression angle exceeded or matched the operative derotation 55% and 18% the time, respectively, though this still leaves 27% uncorrected (9).

It is noteworthy that relatively few studies examining TDO and FDO outcomes in CP focus on foot progression angle, even though this is often a primary factor in the decision for surgery. One possible explanation for this is that FDO and TDO often occur together. Thus, in an observational study, it is difficult to isolate the relative contributions of each procedure. To the best of our knowledge, there has never been an RCT examining the impact of TDO or FDO on foot progression in children with CP. In fact, RCTs studying any orthopedic procedure in CP are rare. There are several explanations for this. First, orthopedic surgery in CP is generally performed as a multi-level intervention. Thus, isolating the effects of individual procedures is a combinatorial challenge. Additionally, although CP is the most common cause of childhood disability worldwide, and has many orthopedic sequelae, surgical volumes at any one center are insufficient for proper RCT design. At the same time, the financial support and incentives required to perform large multi-center studies are simply not available due to the chosen priorities of both public and private funding sources.

In this study we will focus only on foot progression deviations. We will be analyzing retrospective data – and in some cases the primary goal of the derotational osteotomy may not have been correction of foot progression. Regardless, the results will show the effect on foot progression deviations, and thus be useful for our primary goal.

### 1.3 Short-term Foot Progression Outcomes at Our Center

We have observed an unpredictable success rate in correcting foot progression deviations at the initial follow-up evaluation, which is 18 months from baseline on average (Table 1). At our center, in over 2000 limbs of children with CP, 24% of foot progression deviations were worse, 34% were unchanged, 24% were improved or over-corrected, and 18% were fully corrected (within typical limits). These results reflect the effects of derotational osteotomies in combination with other surgeries, natural progression, as well as the decision not to perform surgery. In other words, this is the “*as treated*” effect, reflecting the combination of treatment decisions and treatment effects. It is also important to recognize that changing foot progression may not have been a treatment goal in many cases.

**Table 1.** Historical Outcomes for Foot Progression Deviations.

|  | Worse | Unchanged | Improved | Corrected | Over-Corrected |
| --- | --- | --- | --- | --- | --- |
| **Baseline** |  |  |  |  |  |
| External | 7% | 13% | 7% | 7% | 2% |
| Typical | 14% | 12% | 0% | 0% | 0% |
| Internal | 3% | 9% | 8% | 11% | 7% |
| Total | 24% | 34% | 15% | 18% | 9% |

*Worse* = change $>5^\circ$ away from typical and follow-up outside typical limits,
*Unchanged* = change $<5^\circ$ ,
*Improved* = change $>5d$ towards typical,
*Corrected* = follow-up level within typical limits.
*Over-corrected* = change to opposite deviation but smaller than baseline level

### 1.4 Problems with Observational Studies

Observational results, such as those presented in Table 1, are currently the best evidence we have for understanding the treatment effects of TDO and FDO on foot progression in children with CP. As is widely understood, observational studies are not ideal since they lack the proper controls for effects not caused by the treatment of interest. In the case of foot progression deviations, these concomitant effects include those arising from surgery (e.g., FDO when considering TDO, TDO when considering FDO, foot reconstruction), remodeling of long bones and changes to foot structure over time, possible changes in gait with changes in other pathology (e.g., contracture, spasticity), and maturation of gait pattern with age and walking practice. In the model that we propose below, these other effects will be separated from the effect arising from the treatment of interest (TDO or FDO).

Another important problem with observational studies is that they are inherently susceptible to selection bias. Patients receiving different treatments are not randomized, and thus differ in their characteristics. In addition, patients are generally chosen for treatments based on a doctor’s reasonable belief, based on evidence and experience, that the patient will do well with a treatment, poorly without a treatment, or both. This last element of bias is known as “*targeted selection*” (sometimes called “*The Perfect Doctor Paradox*” in the Rubin causal model framework) and causes important but often unrecognized problems when modeling treatment outcomes in observational studies (10,11).

### 1.5 Causal Inference

An RCT is the gold standard for measuring causal effects, but it is not the only means for obtaining an causal estimate (12). There are many statistical and machine learning methods that can be used to estimate treatment effects (13). All of these rely on adjusting for and regressing on important covariates that determine both treatment assignment and treatment outcome. Recent entries to this inferential arsenal are methods exploiting the Bayesian Additive Regression Tree (BART) framework (14). Stated simply: BART models build a collection of small regression trees that are added together to make a prediction. There is a huge literature describing BART models and their applications to many problems. Of note for this study is the work of Hill et al. who demonstrated the principles by which BART-based models achieve accurate causal predictions (15). This was followed up by the work of Dorie et al. who compared a large number of state-of-the-art causal inference methods on a large set of challenging datasets (16). Dorie’s study showed that BART-based methods performed exceptionally well and provided more accurate and precise treatment predictions than other inferential methods.

### 1.6 Bayesian Causal Forests

A recent addition to the BART-based methods is an approach known as Bayesian Causal Forests (BCF) (11). The BCF method uses a modification of the general BART approach to provide protection against targeted selection and the bias it can introduce (known as regularization induced confounding). The key innovation in the BCF model is to treat the response (predicted outcome) as a sum of a treatment effect (*τ*) plus a concomitant effect from other factors (*μ*). The treatment effect is the object of interest here, while the concomitant effect is the sum of treatments other than the procedure of interest plus effects that arise with age and maturation. Both *τ* and *μ* are assumed to depend on a set of chosen covariates (***x***). So, for example, if the treatment of interest was a TDO, then the concomitant effect would include other surgeries, including FDO, as well covariates like age or sex. It is worth reiterating here that a strength of BART-based approaches is the ability to allow general interactions between covariates, so in the example just given, age and sex could have main effects as well as interactions with each other and with the TDO. A second key modification of BART in the BCF approach is to explicitly include the propensity score (*π*) in the model along with the other chosen covariates. The propensity score is the probability that a particular limb (in this study we will focus on limbs rather than patients) will receive a given treatment. In other words, *π*_*TD0*_(***x***) is the probability of a limb undergoing a TDO, given a set of covariates (***x***) such as age, bimalleolar axis, foot progression angle, etc. The explicit inclusion of the propensity score, along with the separation of *τ* from *μ*, allows BCF to mitigate the effects of sample bias.

### 1.7 Heterogeneous Treatment Effects

Individuals differ from one another in meaningful ways, such as age, sex, comorbidities, prior treatments, and countless other factors. These differences may affect how they respond to a particular treatment. As a result, the average treatment effect may not provide useful guidance for a clinician when evaluating an individual patient. Heterogeneous treatment effects (HTEs) describe the variation of treatment response across individuals. There is an obvious clinical value in identifying covariates related to HTEs, or “*risk factors*”, in the parlance of our times. Knowledge of these risk factors can help clinicians predict how an individual will respond to treatment, leading to better outcomes and guiding patient expectations.

We expect a pronounced HTE for derotational osteotomies depending on preoperative deformity. This is because there is a natural dose dependence on preoperative deformity magnitude and direction; larger preoperative torsional deformity leads to more foot progression deviation, which, in turn, leads to larger operative derotational magnitudes, and thus larger treatment effects. It is worth noting that it would be ideal to include the amount of derotation as a covariate, since this would likely be a predictor of treatment effect. Unfortunately, the amount of derotation is not routinely reported, so we will estimate the change in torsion from pre- and postoperative physical examination-based measures. It is important to remember that this measured change is not necessarily equal to the surgical correction since it includes the effects of both surgical derotation and bony remodeling. As a result, we will not include this estimate as a covariate in the model. We will, however, examine HTEs with respect to this measured change in torsion.

### 1.8 Derotational Osteotomies as a Model

Foot progression deviations are a common and meaningful problem, and derotational osteotomies are the primary tool for treating them. Additionally, studying foot progression and derotational osteotomies also allows us to fulfill our secondary goal of demonstrating the value of BCF for both identifying causal effects and analyzing HTEs since (i) both TDO and FDO affect foot progression deviation, (ii) TDO and FDO often occur together, making estimates of their causal effect difficult, (iii) we expect confounding due to other treatments, such as foot surgery, which frequently occurs alongside TDO and FDO, (iv) we expect confounding due to bony remodeling, and this confounding is age and sex dependent, and (v) there are sound mechanistic bases for the sources of HTEs, and we can test whether the treatment effect estimated by BCF identifies these risk factors.

## 2 Methods

This study involved data previously collected for clinical purposes. Patients gave written consent for the use of their medical records in research and publication when the clinical services occurred. Data were recorded so that individuals could not be identified directly or through identifiers. The University of Minnesota Institutional Review Board (IRB) ruled that the study (STUDY00012420) was not research involving human subjects as defined by the Department of Health and Human Services and Food and Drug Administration regulations. To arrive at this determination, the IRB used “WORKSHEET: Human Research (HRP-310).” (17).

### 2.1 Participants and Data

We searched our database between January 2006 and December 2020. Limbs of patients meeting the following criteria were selected for analysis:

- Diagnosis of CP.
- Less than 18 years old at baseline gait evaluation.
- Baseline and follow-up gait analysis within 3 years of each other.
  - To estimate treatment effects for TDO and FDO we seek a dataset including limbs that underwent TDO or FDO, other surgeries besides TDO or FDO, or no treatment at all. Thus we place no criteria regarding intervening treatments.
- Both kinematics and physical examination data present.
  - Missing data for categorical measures (e.g., manual muscle strength score) were treated as a unique category (“missing”) rather than being imputed or dropped. Limbs with missing scale data (e.g., maximum passive internal hip rotation) were dropped from the analysis.
- Swing phase knee varus-valgus range-of-motion <15° to ensure rotational kinematic profile quality (18).

### 2.2 Gait and Clinical Examination Measures

Three-dimensional gait kinematics were measured at baseline and follow-up using three to five barefoot over-ground walking trials collected at a self-selected speed. The motion analysis laboratory used modern, three-dimensional gait analysis equipment and methodology and employed highly experienced staff. Kinematics were computed using a modification of the Vicon Plug-in-Gait model (Vicon Motion Systems Ltd, UK) with hip centers and knee axes identified using functional methods, and malleoli identified using virtual markers (19,20). Foot progression angle was defined by projecting a line from the ankle center to a point between the heads of the second and third metatarsals onto the laboratory floor and computing the angle with respect to the direction of walking.

Physical examinations were performed by a licensed physical therapist at baseline and follow-up. Spasticity was scored using the modified Ashworth scale (21). Strength was estimated from a manual muscle test (22). Static selective motor control at various levels was graded as absent, diminished, or typical. Range-of-motion was assessed passively using a hand-held goniometer. Prior surgical history was extracted from our clinical database.

Propensity scores for TDO and FDO were computed from a separate BART model using the `bartMachine` package in R (23). Propensity score modeling is not the focus of this paper, and many good methods exist for estimating propensity scores (24). The performance of propensity models used in this study is included for reference (Figure 1).

**Figure 1.**
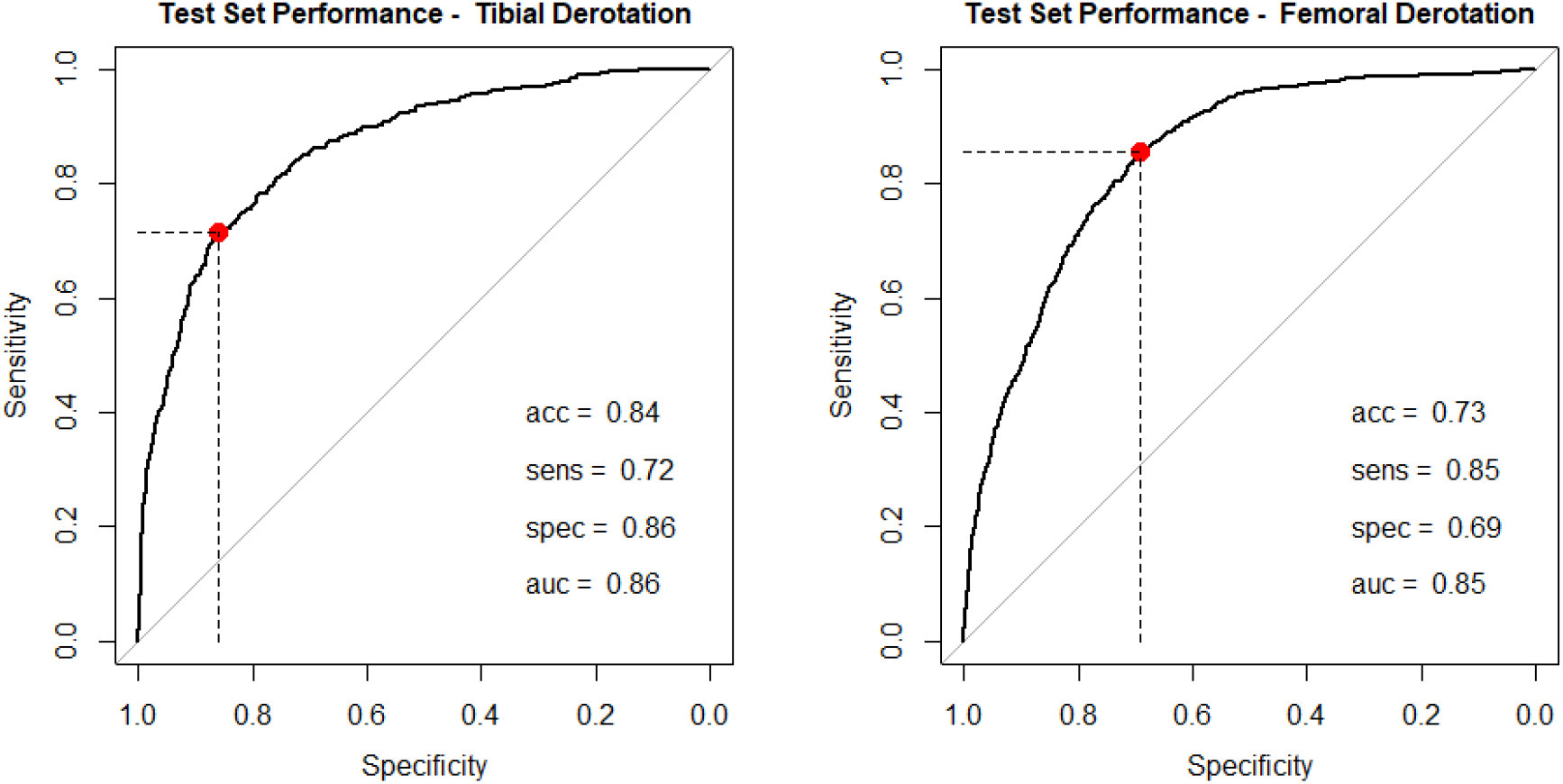
Propensity score performance for TDO and FDO on independent test data. The reported metrics represent the levels at the chosen threshold (red dot and dashed lines).

### 2.3 Outcome measure and Covariates

Our outcome of interest was the change in mean stance-phase foot progression angle (follow-up minus baseline).

The covariates ***x*** were chosen based on clinical experience. We included variables commonly used during discussions of treatments for patients. The list of covariates is as follows:

- **Anthropometry**: age, height, leg length, sex.
- **Time and Distance parameters**: foot-off, opposite foot-off, opposite foot contact, dimensionless speed, cadence, and step length (25).
- **Neurological**: *<u>Spasticity</u>* (Modified Ashworth Scores for hip adductors, hamstrings, plantarflexors, rectus femoris), *<u>Strength</u>* (Manual Muscle Test Grades for hip abductors, knee extensors and flexors, plantarflexors), *<u>Static selective motor control</u>* (Absent, Diminished, or Typical for hip abductors, knee extensors and flexors, plantarflexors).
- **Range of motion**: Ankle dorsiflexion (knee extended and flexed), knee extension, popliteal angle, hip abduction (knee extended and flexed), hip rotation (external and internal).
- **Bony alignment**: bimalleolar axis angle (surrogate for tibial torsion) and trochanteric prominence test angle (surrogate for femoral anteversion).
- **Gait:** *<u>Angles</u>*: sagittal, coronal, and transverse plane for the pelvis, hip, and knee, sagittal plane ankle, transverse plane foot, *<u>Measures</u>*: Level and timing of maximum and minimum during initial contact, stance, foot-off, swing, and overall, level of mean and range of motion during stance, swing, and overall.
- **Surgery**: Prior and concomitant.
- **Propensity score**: probability of receiving a TDO or FDO.

### 2.4 Model Details

All computations were performed in R (26). We used the BCF package with default settings (11).

## 4 Results

### 4.1 Limb Characteristics

We analyzed 2302 limbs from 1279 baseline-to-follow-up pairs of visits for 774 individuals (Table 2). Note that the number of limbs is not exactly twice the number of visits due to removal of observations with missing scale data or significant knee varus/valgus artefact. Of the limbs analyzed, 147 had a TDO without FDO, 282 had an FDO without TDO, 190 had both a TDO and an FDO, and 1683 had neither a TDO nor an FDO.

**Table 2.** Baseline Limb Characteristics

|  | No TDO<br>No FDO | Yes TDO<br>No FDO | No TDO<br>Yes FDO | Yes TDO<br>Yes FDO |
| --- | --- | --- | --- | --- |
| Number of Limbs | 1683 | 147 | 282 | 190 |
| Sex, Percent Male | 57 | 71 | 56 | 59 |
| Age | 8.7 (3.4) | 10.6 (2.9) | 9.5 (3.0) | 9.3 (2.9) |
| Bimalleolar Axis Angle | 10.9 (11.2) | 19.1 (15.8) | 13.6 (9.5) | 16.5 (16.2) |
| Trochanteric Prominence Angle | 34.4 (16.3) | 30.8 (14.8) | 47.9 (12.8) | 48.2 (11.7) |
| Passive Hip Internal Rotation | 58.9 (14.1) | 56.5 (13.6) | 69.7 (10.9) | 69.7 (10.5) |
| Passive Hip External Rotation | 36.8 (12.7) | 38.9 (13.1) | 29.6 (13.5) | 29.6 (13.0) |
| Mean Stance Pelvic Rotation | 0.5 (7.4) | 0.5 (7.1) | -1.6 (7.2) | -1.6 (7.3) |
| Mean Stance Knee Rotation | -9.3 (14.0) | -20.8 (19.5) | -11.1 (10.5) | -14.1 (19.7) |
| Mean Stance Hip Rotation | 1.7 (10.8) | 1.4 (10.1) | 8.2 (11.4) | 8.2 (10.0) |
| Mean Stance Foot Progression | -4.7 (15.4) | -15.4 (21.1) | -1.1 (15.2) | -6.5 (20.6) |

*All measurements in degrees except Age (years) and Sex (percent)*
*All values mean (sd) except Sex expressed as number (percent) in treatment sub-group*

### 4.2 Model Performance

#### Overall Performance

To put the BCF results in context we compared the BCF predictions to five other models:

- **Corrected** – assume mean stance-phase foot progression reaches the typical level (−5.8°) at follow-up.
- **Unchanged** – assume mean stance-phase foot progression does not change from baseline to follow-up.
- **Mean Response** – assume the change in mean stance-phase foot progression of each limb equals the mean historical change for all limbs (−3.7°).
- **Linear** – predict the change in mean stance-phase foot progression with a linear model based on the covariates ***x***.
- **BART** – predict the change in mean stance-phase foot progression using a standard (non-causal) BART model based on the covariates ***x***.

The BCF model was able to predict follow-up foot progression with an overall RMSE accuracy of 7.8° (Figure 2). This was the best performance among the six models examined, with an error half as large as the corrected, unchanged, and mean response predictions, and marginally better than an ordinary BART model.

**Figure 2.**
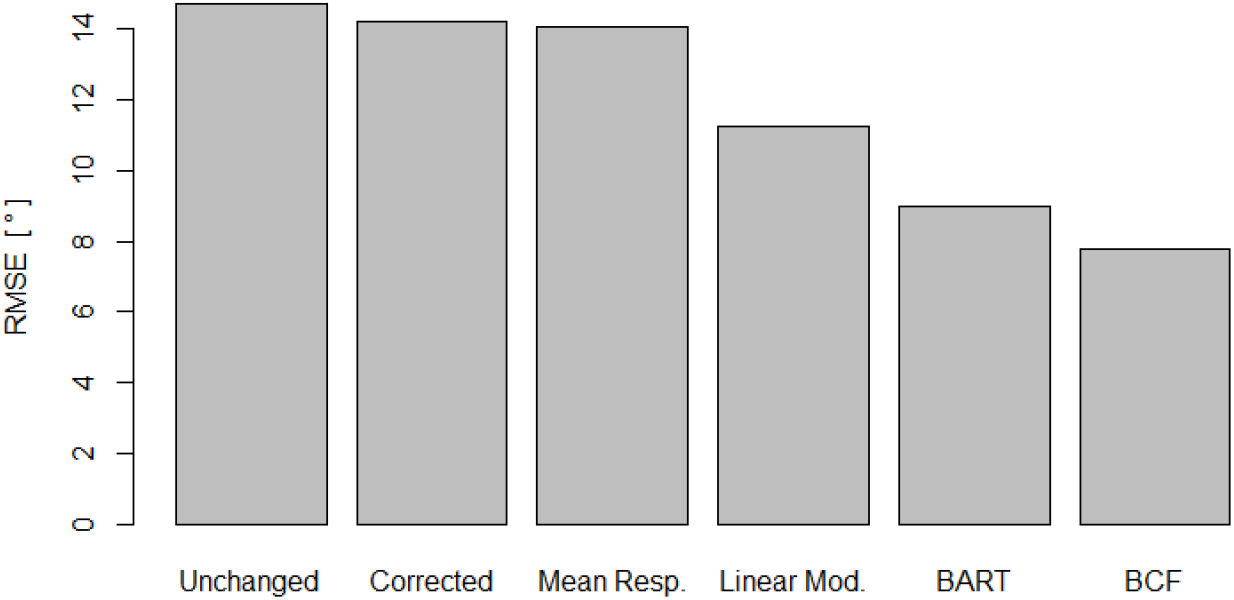
Predictive accuracy of various models. The BCF model for TDO outperformed the other models, including (non-causal) BART. Note that BCF model for FDO performed nearly identically.

The BCF model showed balanced performance across the four treatment groups defined by TDO and FDO status (Figure 3). This suggests the model is not dominated by predicting one group (TDO only, FDO only, TDO and FDO, Neither) or one component of outcome (*μ* or *τ*) at the expense of other groups or components. This is an important finding since the group sizes are unbalanced, and superb performance in a large group (e.g., neither TDO nor FDO, N = 1683), could have (but did not) mask poor performance in a smaller group (e.g. TDO without FDO, N = 147).

**Figure 3.**
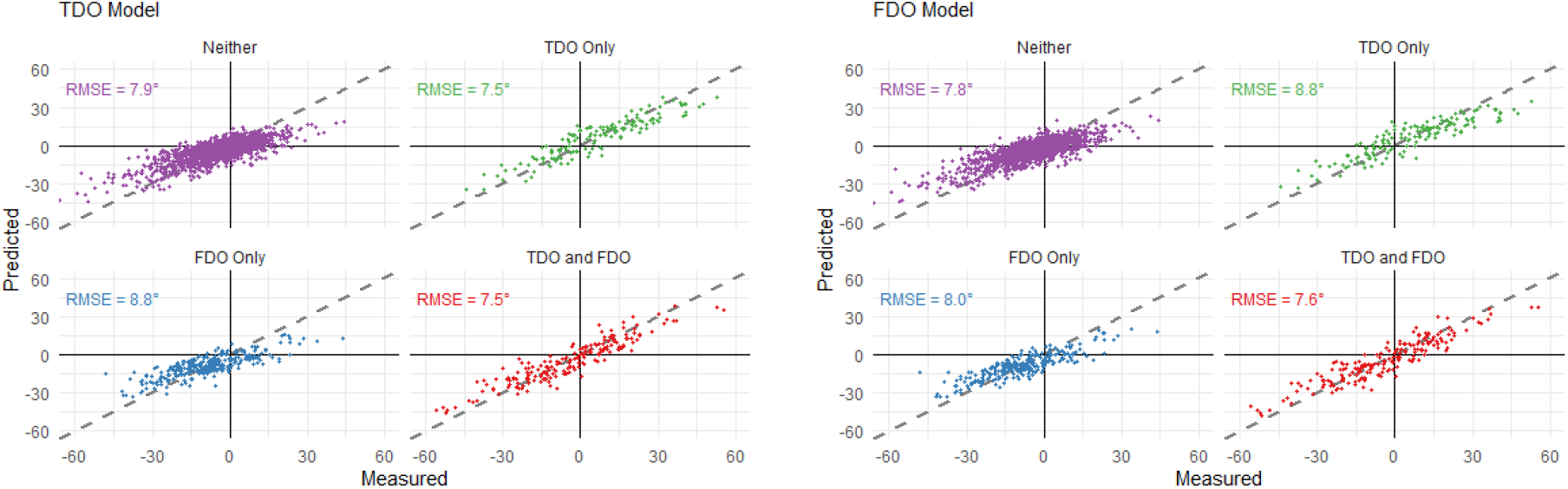
Performance of BCF model across treatment groups. Perfect prediction is shown by the dashed parity line. Prediction accuracy was well balanced across the treatment groups.

### 4.3 Treatment Effects

#### Marginal Distribution of Treatment Effects

Marginal distributions of treatment effect (*τ*) and concomitant effects (*μ*) from FDO and TDO show the existence of significant HTEs, especially for TDO (Figure 4). We observed a bi-modal distribution of *τ*_*TDO*_, reflecting the fact that tibial torsion deformities occur in both directions (internal and external), and hence corrections occur in both directions as well. The lack of density around 0 for the *τ*_*TDO*_ suggests that the surgery is not prescribed unless a significant derotation is needed. In contrast, the distribution of *τ*_*FDO*_ was unimodal and externalizing, reflecting the dominance of excessive femoral anteversion in children with CP.

**Figure 4.**
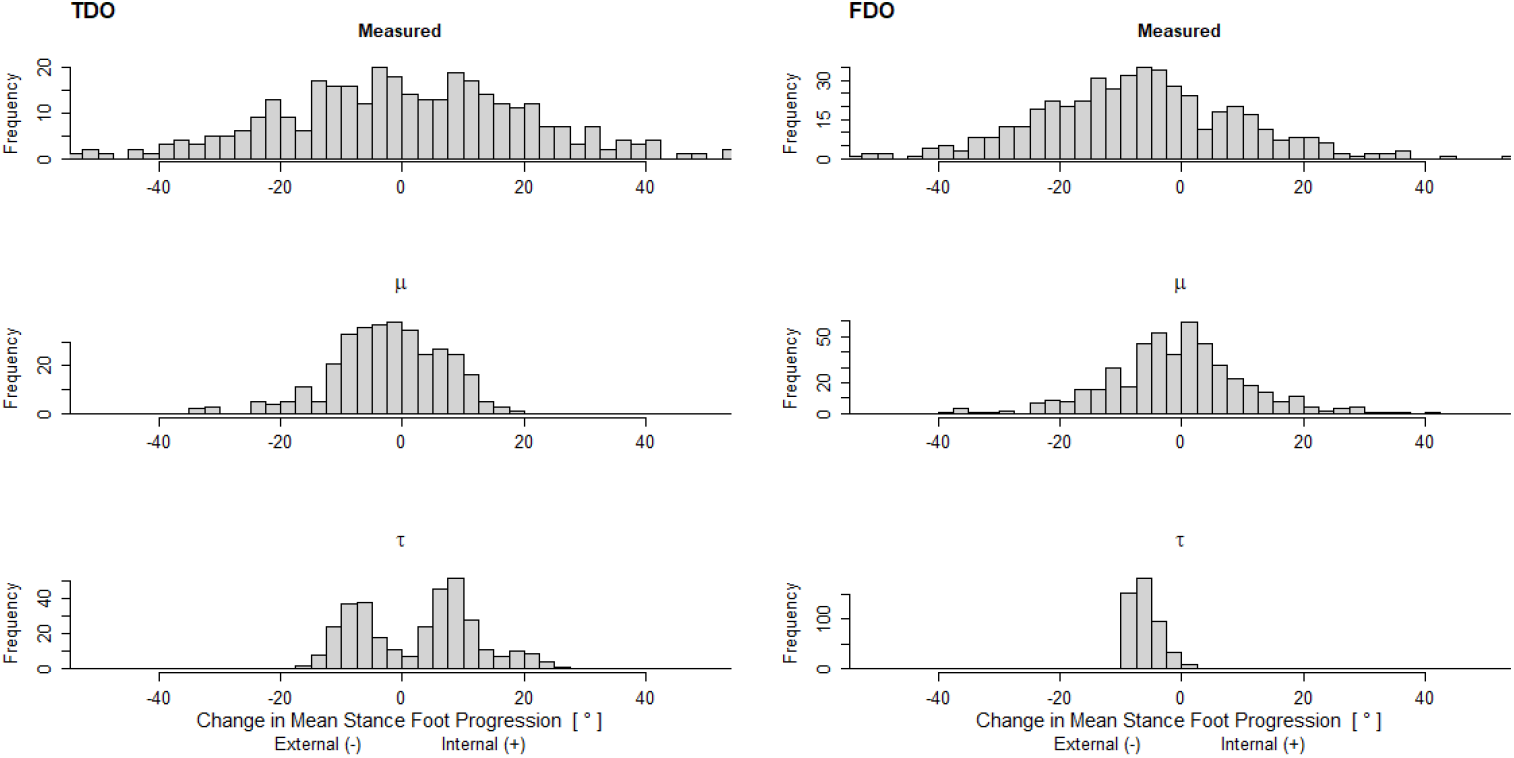
Results for TDO (left) and FDO (right). The *measured* outcome (top), *modeled* concomitant effects *μ* (middle), and *modeled* treatment effect *τ* (bottom) are shown. Note that the TDO treatment effect has a bimodal distribution since children with CP exhibit both external and internal tibial torsion deformity. In contrast, the FDO treatment effect is almost exclusively externalizing, consistent with the fact that excessive anteversion is the overwhelming direction of the deformity in this population.

#### Treatment Effect Estimate Precision

An important aspect of BCF is that, as a Bayesian method, it produces a posterior probability distribution for all parameters, rather than simply a point estimate. This means that we have the full distribution for *τ* (and *μ*) for each limb in the data, allowing us to calculate limb-by-limb uncertainty estimates. We extracted the posterior distribution of *τ* for each limb and calculated this distribution’s standard deviation as a measure of prediction precision. On average, the standard deviation of the treatment effect *τ*_*TDO*_ was 4° and *τ*_*FDO*_ was 2.7°, with 95% of treatment effect standard deviations falling below 6° (Figure 5). In other words, on average, the error (standard deviation) of the predicted treatment effect for an individual limb was around 3° - 4°.

**Figure 5.**
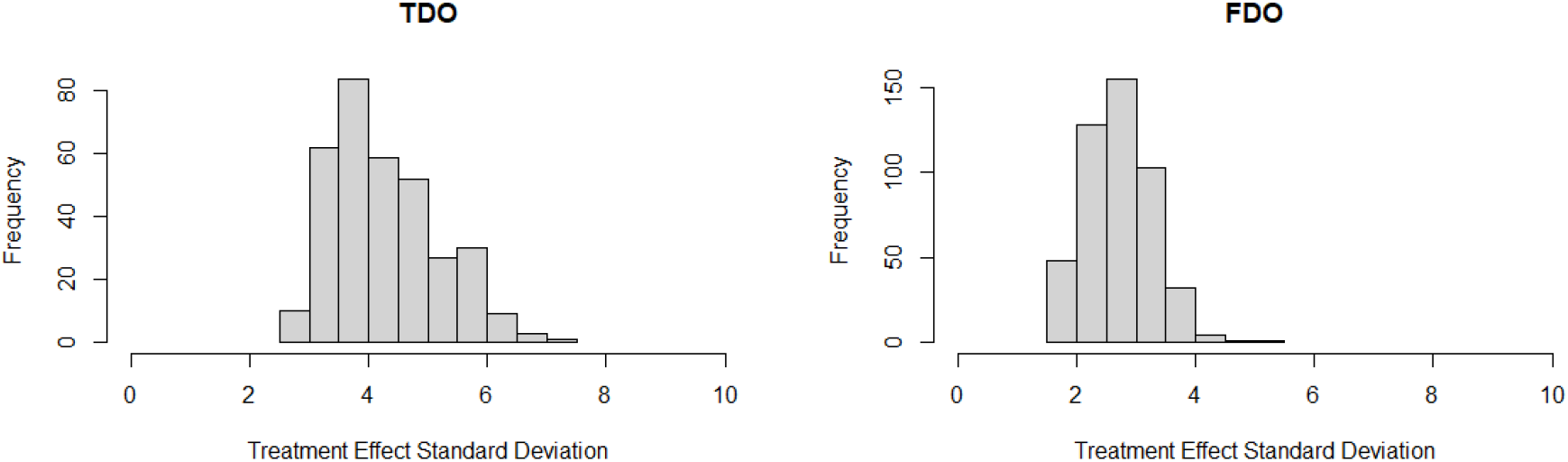
The histograms show the standard deviation of the treatment effect estimates for individual limbs. Median precision was 4.1° and 2.7° for the TDO model (left) and FDO model (right), respectively.

#### Treatment effect relative to total effect

The ratio of treatment effect to total effect varied significantly across the study sample (Figure 6). In the case of TDO, the variation in this ratio came from both the treatment effect (*τ*) and the concomitant effect (*μ*). In the FDO, the treatment effect was relatively constant, but the concomitant effect varied significantly. Recall that the concomitant effect includes bony remodeling, other treatments, age, maturation, and walking practice. Note that in 56% of limbs receiving a TDO the concomitant effects include those arising from an FDO, while in 40% of limbs receiving an FDO the concomitant effects include those arising from a TDO.

**Figure 6.**
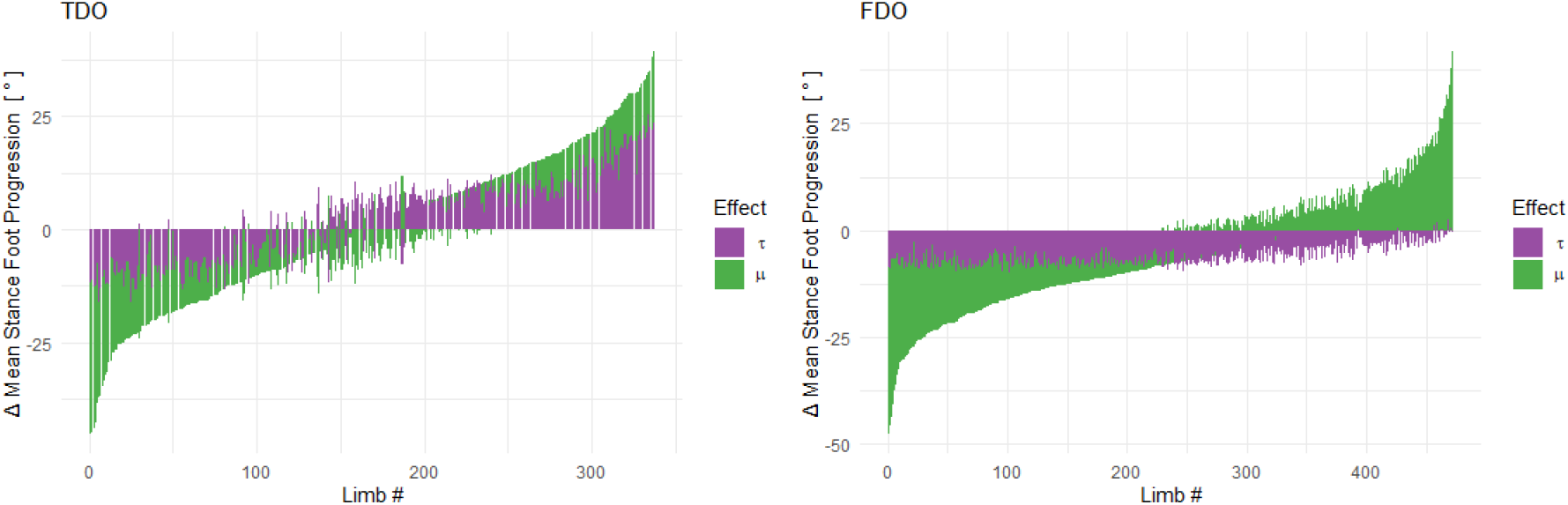
Treatment and concomitant effect per limb for TDO (left) and FDO (right). The plots are arranged from largest negative to largest positive total effect. The purple bar shows the treatment effect (*τ*) and the green bar the concomitant effect (*μ*). Note that the treatment effect for TDO shows significant heterogeneity and bidirectionality, while the treatment effect for FDO is largely homogeneous and unidirectional.

#### Heterogeneous Treatment Effects

The effect of a TDO varied substantially with preoperative tibial torsion and foot progression (Figure 7 - *top row*). The effect of an FDO was nearly constant with respect to preoperative anteversion and foot progression (Figure 7 - *bottom row*).

**Figure 7.**
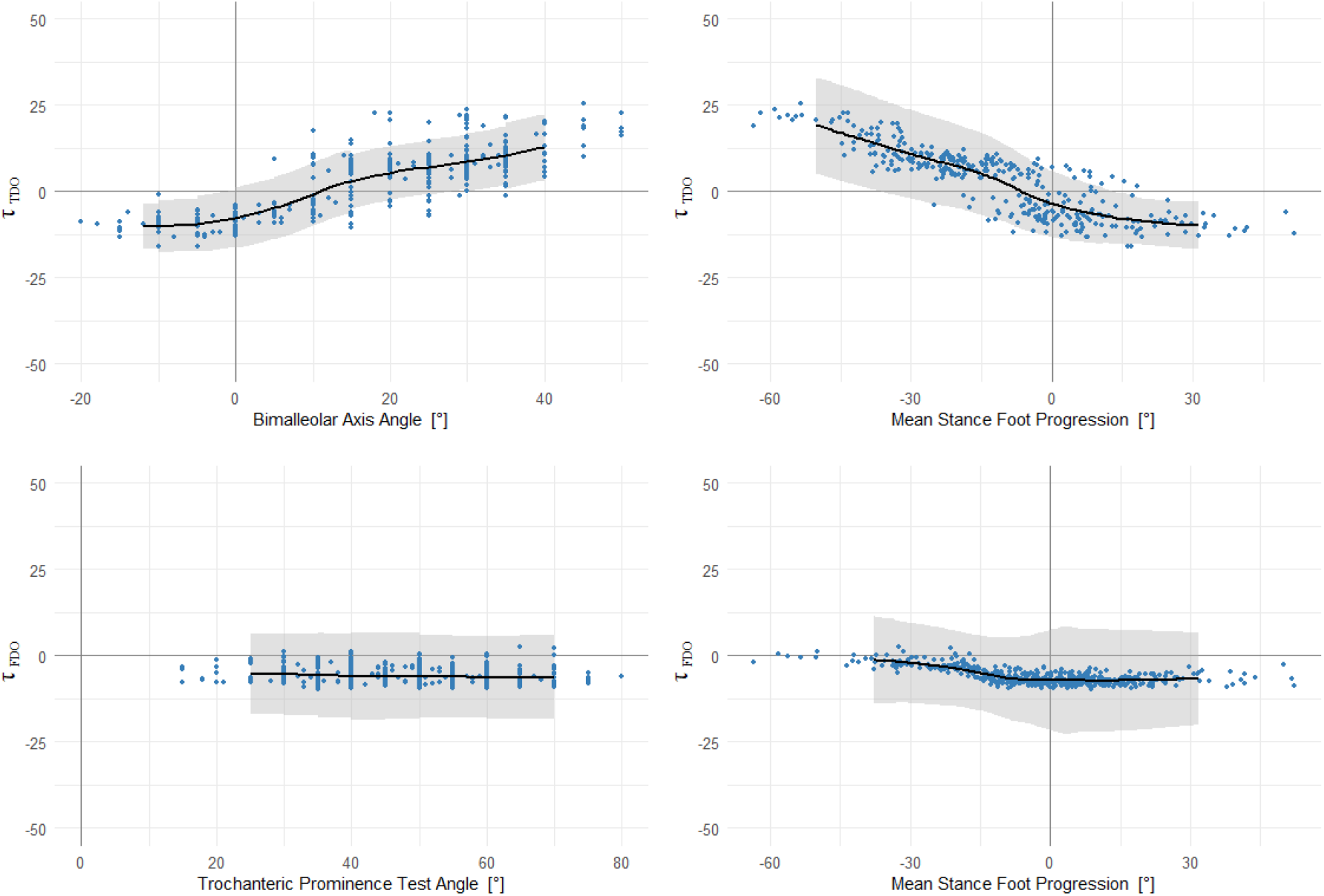
(*top row*) Treatment effect for TDO as a function of estimated preoperative tibial torsion (left) and mean stance foot progression (right). (*bottom row*) Treatment effect for FDO as a function of estimated f preoperative anteversion (left) and mean stance foot progression (right). The grey band is ±1 sd and the y-axis scale is 90% of the total effect range observed in the measured data.

The treatment effect for TDO varied with the estimated amount of tibial torsion change (surgical correction + bony remodeling) (Figure 8 - left), while the treatment effect for FDO did not vary with the estimated amount of femoral anteversion change (Figure 8 - right).

**Figure 8.**
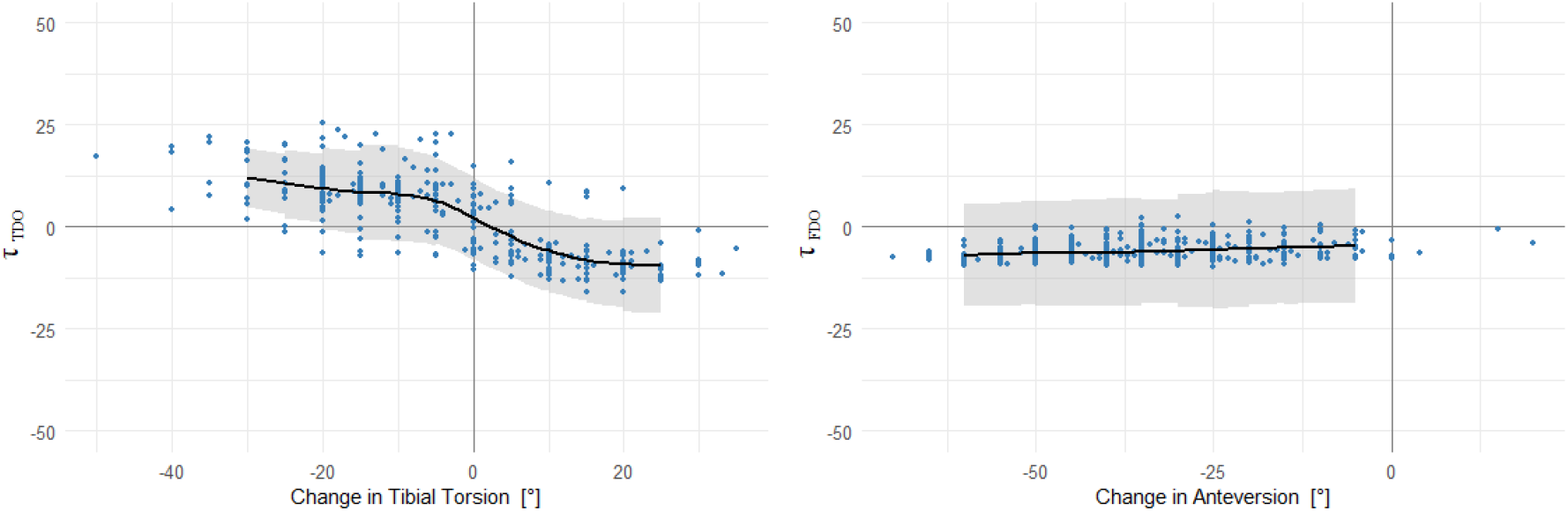
Treatment effect for TDO (left) and FDO (right) as a function of estimated amount of operative correction. The grey band is ±1 sd and the y-axis scale is 90% of the total effect is 90% of the total effect range observed in the measured data.

#### Effect Specificity

We did not expect the effect of a TDO to depend on the change in anteversion, nor did we expect the effect of an FDO to depend on the change in tibial torsion. This was confirmed in our results (Figure 9). This effect specificity was consistent with the well understood mechanism of correction through bony derotational and with our proposal that the BCF method is not conflating treatment effects with concomitant effects.

**Figure 9.**
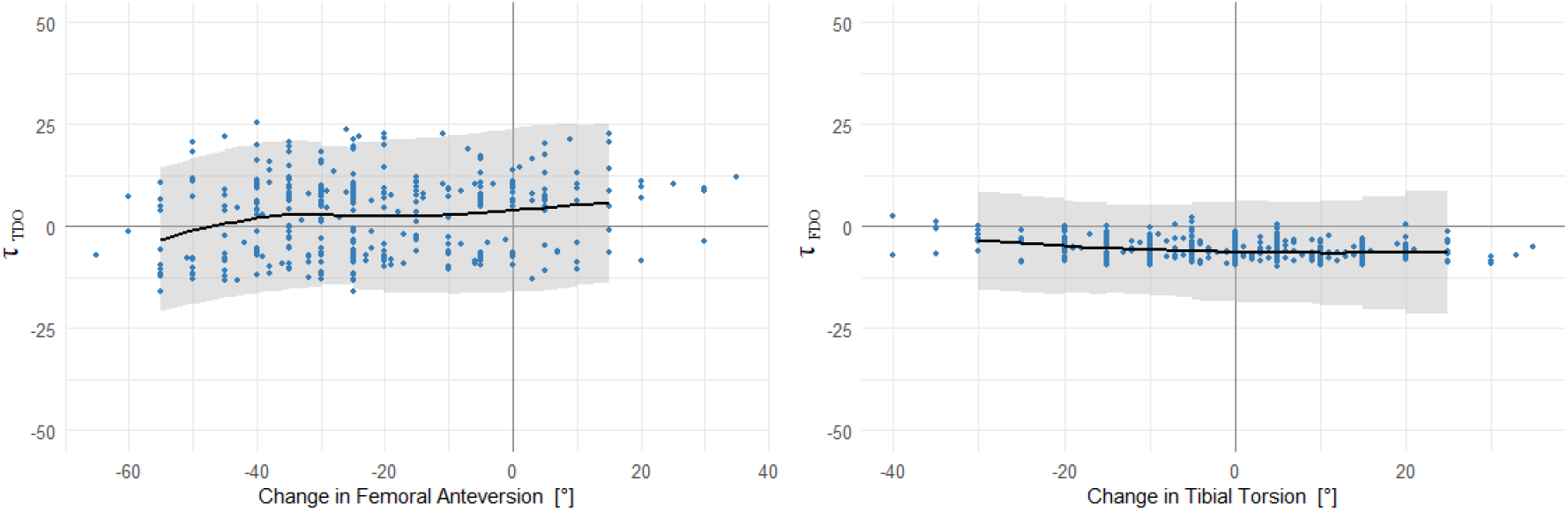
The treatment effect of TDO does not vary with change in femoral anteversion (left) and the treatment effect of FDO does not vary with the change in tibial torsion (right). This confirms that the BCF model is not conflating treatment and concomitant effects. The grey band is ±1 sd and the y-axis scale is 90% of the total effect range observed in the observational data.

## 5 Discussion

We estimated causal treatment effects of TDO and FDO on mean stance-phase foot progression, identified clinically meaningful HTEs, and demonstrated the use of BCF for understanding treatments and outcomes in children with CP. It is important to note that, as with any causal inference problem, we lack a gold standard against which we can judge our model. We will instead rely on prior literature, observed marginal distributions, and clinical understanding of the anatomy and surgeries for assessing the quality of the estimates.

### 5.1 TDO results

We found that, for a TDO, the model identified distinct directions of effect (external and internal), consistent with the bi-directional tibial torsion deformity commonly found in children with CP. We found a monotonic, relatively linear dependence of TDO treatment effect on both preoperative torsion and preoperative foot progression, with zero-crossings at almost exactly the typical values. Furthermore, we found a strong HTE with respect to the estimated change in tibial torsion (derotational + remodeling). These causal trends were consistent with the marginal effects, though attenuated in magnitude (Figure 10).

**Figure 10.**
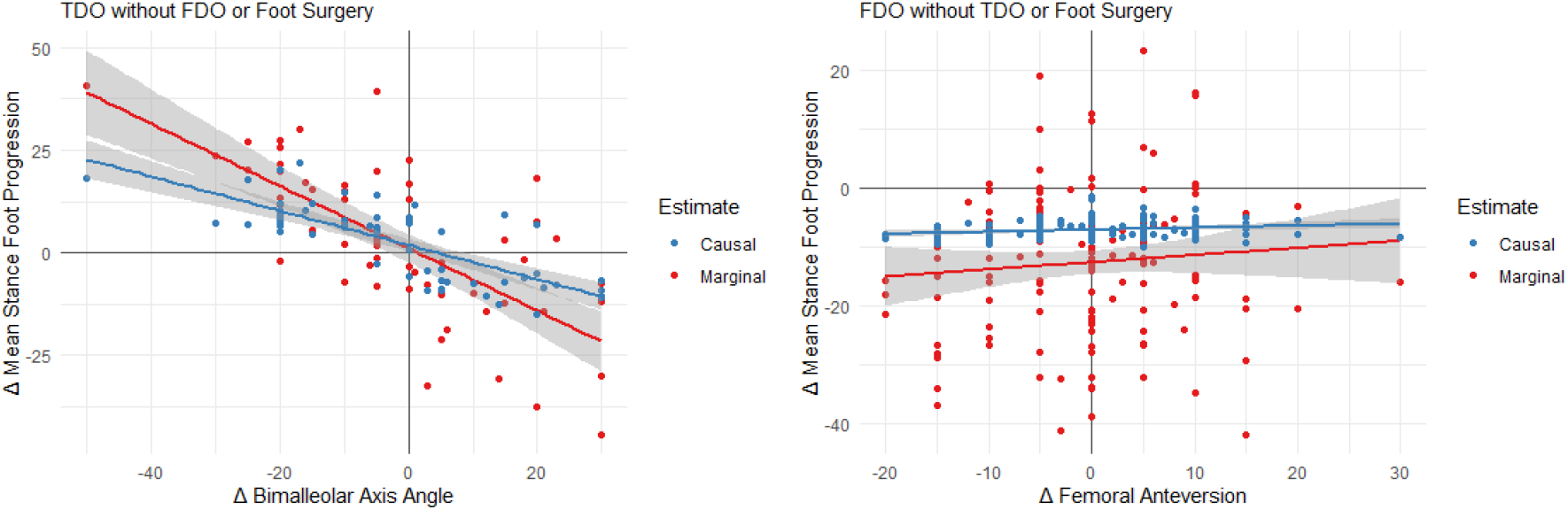
Causal and marginal outcome distributions. The plots show change in mean stance foot progression versus change in bimalleolar axis angle (left) and femoral anteversion (right) for limbs undergoing a TDO without FDO or foot surgery (left) or FDO without TDO or foot surgery (right). For TDO, both the BCF model estimate of *τ*_*TDO*_ and the marginal distribution were strongly dependent on the change in bimalleolar axis angle. For FDO, both the BCF model estimate of *τ*_*FDO*_ and the marginal distribution were independent of the estimated amount of anteversion change. The magnitude difference between marginal and causal estimates is likely due to residual concomitant effects.

The treatment effect for TDO does not scale one-to-one with the estimated change in tibial torsion (linear model slope ∼0.5). The discrepancy could arise from several sources, including compensation at other levels, recurrence of deformity, and bias errors in our estimate of tibial torsion. This last possibility is particularly intriguing considering research that suggest imaging-based tibial torsion assessment following TDO does not match intraoperative documentation (27). The geometry of the distal tibia, the details of operative correction (e.g., fibula sparing), and the radiographic or physical examination-based estimates of tibial torsion all suggest discrepancies between operative correction definitions and radiographic or physical examination-based measures.

### 5.2 FDO Results

We found that the treatment effect for an FDO was consistently externalizing. This is consistent with the fact that excess femoral anteversion dominates as the direction of deformity in children with CP. Thus, almost all femoral derotations are externalizing, even most of those not intended to treat foot progression (e.g., used to correct hip deformity). We found a nearly constant FDO treatment effect, with little dependence on preoperative anteversion, preoperative foot progression, or change in femoral anteversion following FDO. These finding are surprising, since we expected that the amount of baseline deformity and intraoperative correction would influence the outcome. However, further inspection showed that the model results are consistent with the marginal distribution of limbs that underwent an FDO with neither TDO nor foot surgery (Figure 10). Thus, the counterintuitive revelation from the BCF model alerted us to a fact that we had previously overlooked. The ability of the BCF to identify this novel and important result points to the value of causal modeling, and the power of the BCF method to discern treatment effects from observational data.

### 5.3 Effect Specificity

In addition to observing expected heterogeneity with preoperative deformity level, we also found that heterogeneity was absent where it should be absent. The large TDO effect showed virtually no dependence on change in anteversion, and, likewise, the moderate FDO effect showed no dependence on change in tibial torsion. In other words, even though the two treatments occurred simultaneously in 190 limbs, there was no “*leakage*” artefact in the identified treatment effect or its dependency on clinical variables. This is further evidence, albeit indirect, that the BCF algorithm was properly identifying causal treatment effects.

### 5.4 Limitations

A fundamental problem with causal inference models is that there is no “*gold-standard*” that can be used to confirm or refute model results. Consequently, we must rely on indirect evidence to support our claims. The indirect evidence in this study supports our claims of causal identification, and we did not find any results contrary to the claim that the model worked. However, further experiments (e.g., prospective predictions) are needed to test the model performance.

As with any retrospective study there is a possibility of selection bias. In this study, we only evaluated patients referred for gait analysis. This sample may not be representative of the entire population of ambulatory CP. Perhaps these children are more severe or less severe or showing signs of worsening or showing signs of improvement, and these are the reasons they were sent for three-dimensional gait analysis. Further complicating matters is that our study only involves children with a pair of gait analysis less than three years apart. We know that a moderate proportion of children seen in our motion analysis center for a baseline evaluation do not return for follow-up, regardless of whether they received treatment. It is possible that the missing children responded differently than those who returned.

The covariates were chosen subjectively, based on clinical experience and guidance from the literature. It is possible that key covariates that could have improved the model were missed. The covariates we chose were extensive and comprised those used daily for making these treatment decisions. Some of our covariates have large experimental uncertainties associated with them. Physical exam measures of torsional deformities are plagued by imprecision and bias. There can also be significant errors in transverse plane kinematics due to methodological errors and soft-tissue artefacts. We have procedures in place to mitigate these as much as possible, including the use of functional model calibration to identify knee axis, resulting in the removal of data that showed significant knee axis misalignment (17% of observations). In some sense, data quality problems further support the use of BCF or other strongly regularized algorithms. Such algorithms are inherently skilled at rejecting irrelevant data and not being swayed by noise. The BCF algorithm achieves this through its separation of the treatment and concomitant effects and imposition of strong regularization on the treatment effect. We did not find any HTEs in FDO, and this was supported by the marginal distribution analysis. However, it is worth noting that parameter choices for the BCF model affect the likelihood of detecting HTEs. We have used the default parameters, which have been shown to work well over a wide range of problems.

### 5.5 Future Work

The change in tibial torsion or femoral anteversion by surgery is the actual causal variable of interest. The clinical question we really want to answer is “*how much should a given bone be derotated to achieve a desired change in foot progression?*”. Unfortunately, the amount of derotation is not explicit in model since it is not routinely documented. The dichotomization of the treatment to yes or no TDO/FDO, rather than TDO of 23° internal or FDO of 18° external, is an artificial construct. It would be useful to build a model with amount of derotational as a causal variable and use the model results to guide surgery dose for preoperative planning.

It is notable that the programming required to change the outcome variable studied is trivial; a single variable in a single line of code. For example, in the case of TDO and FDO we might be interested in how the surgeries affect the knee varus moment, or walking speed, or any number of other parameters. Examining a different treatment is also trivial, and is precisely what we do in this study, swapping between TDO and FDO with the change of a single letter, T → F, in our code. This highlights the power of analyzing treatments in the BCF framework.

Testing the model’s performance in a prospective study would be a valuable next step. Since this is a model with no ground truth comparison, implementation details are important and use in a clinical setting must be accompanied by clear and understandable disclosures of model assumptions and limitations.

## Data Availability

**Data availability**
The data are not publicly available due to them containing information that could compromise research participant privacy or consent. Explicit consent to release data was not obtained from the patients, and data were collected up to 15 years ago. Thus, the vast majority of patients cannot be asked to provide their consent for release of their data. The data that support the findings of this study are available from the corresponding author (MHS) upon reasonable request and subject to data sharing agreements.

